# Randomized, double-blind, sham-controlled trial of an intraoral photobiomodulation device for oral mucositis due to radiotherapy for head and neck cancer

**DOI:** 10.64898/2026.02.26.26347195

**Authors:** Kenneth Hu, Pratish Shah, M. Connie Nguyen, Cornelia McCluskey, Anne Kane, Roger Ove, Christopher D. Willey, Sanford Katz, Omkar Marathe, Sasha Valentin, Jennifer Frustino, Alessandro Villa, Sharon Spencer, Catherine Holtzapfel, Nathaniel Treister, Rajesh V. Lalla

**Author notes:** Corresponding Author: Kenneth Hu, MD, Professor, Department of Radiation Oncology and Department of Otolaryngology-Head and Neck Surgery, NYU Grossman School of Medicine, Co-Director, Head and Neck Center, Department of Radiation Oncology, NYU Langone Health, New York, NY, USA. Dr. Kane is now at the University of Alabama at Birmingham, Birmingham, AL, USA. Clinical Trial Registration Number: NCT03972527. **Author contributions** Conceptualization: R.L. and K.H.; formal investigation, data acquisition and interpretation: K.H., P.S., M.C.N, C.M., A.K., R.O., C.W., S.K., O.M., S.V., J.F., A.V., S.S., and C.H.; writing – original draft preparation: K.H. and R.L.; writing – review and editing: P.S., M.C.N, C.M., A.K., R.O., C.D.W., S.K., O.M., S.V., J.F., A.V., S.S., C.H., and N.T. All authors read and approved the final manuscript. **Funding** Funding for this study, including data collection, monitoring, statistical analysis and medical writing, was provided by MuReva Phototherapy Inc. (Brecksville, Ohio, USA). **Declarations**. **Ethics approval** This study was conducted in full compliance with FDA regulations, ICH guidelines for GCP, and in accordance with ethical principles that have their origins in the Declaration of Helsinki. The study was approved by the respective Institutional Review Boards (WCG IRB, NYU Langone Health IRB, and MetroHealth IRB). **Consent to participate** All participants signed an informed consent to participate in the study.

## Abstract

**Purpose:** This study evaluated the safety and effectiveness of an intraoral light-emitting diode (LED)-based photobiomodulation (PBM) device to reduce the incidence and severity of oral mucositis (OM) from intensity modulated radiation therapy (IMRT) for head and neck cancer (HNC).

**Methods:** This randomized, double-blind, sham-controlled trial enrolled patients with HNC undergoing high-dose IMRT over 6-8 weeks, with or without concurrent chemotherapy. Participants received daily 10-minute PBM or sham treatments immediately before IMRT sessions. Assessments were conducted at baseline, daily and weekly during IMRT, and two weeks post-IMRT.

**Results:** Eighty-five participants (42 PBM; 43 sham) were enrolled across 12 US sites. No device-related adverse events were observed, and 99.5% of initiated sessions were completed. In the intent-to-treat population, severe OM (WHO Grade ≥3) incidence was significantly lower with PBM across six weeks of IMRT (36.8% vs 57.1%; *p* = 0.046) and at two weeks post-treatment (10.8% vs 36.4%; *p* = 0.042). In the per-protocol population, the PBM arm reported significantly greater taste preservation (*p* = 0.034), lower increases in mouth/throat soreness (*p* = 0.029) and throat pain (*p* = 0.028) and needed fewer feeding tube placements (*p* = 0.073) than the control arm.

**Conclusion:** Daily intraoral PBM therapy using an LED-based device was safe, well tolerated, and significantly reduced the incidence of severe OM and associated complications in HNC patients undergoing IMRT with or without concurrent chemotherapy. These findings align with guidelines recommending daily intraoral PBM therapy for preventing cancer therapy-related OM, a dose-limiting toxicity for which effective preventive interventions are needed.

**Trial Registration:** ClinicalTrials.gov Registration Number NCT03972527. Registered on June 3, 2019.

**Concise Summary:** Daily intraoral PBM therapy using an LED-based device was safe, well tolerated, and significantly reduced the incidence of severe OM and associated complications in HNC patients undergoing IMRT with or without concurrent chemotherapy. These findings align with guidelines recommending daily intraoral PBM therapy for preventing cancer therapy-related OM, a dose-limiting toxicity for which effective preventive interventions are needed.

## Introduction

Oral Mucositis (OM) is a common and debilitating toxicity of radiation therapy (RT). While advances in treatment planning and delivery have improved precision, acute mucosal injury remains a persistent clinical challenge. Among patients receiving curative RT for head and neck cancer (HNC), OM occurs in over 90% of cases and approximately two-thirds develop clinically severe OM that prevents oral intake of solid food [1].

Oral mucositis is a treatment-related inflammatory condition characterized by erythema, ulceration, and breakdown of the oral mucosa. OM-related ulcerations can cause severe pain, increase susceptibility to secondary infection, and impair swallowing, taste, and salivary flow, collectively contributing to malnutrition and reduced quality of life [2]. Severe OM represents a dose-limiting toxicity that contributes to unplanned treatment interruptions, hospitalizations, and increased healthcare utilization and costs [1], [2], [3], [4], [5], [6], [7]. Notably, contemporary strategies such as radiation dose de-escalation in the Human Papillomavirus (HPV)+ oropharynx population [8], [9] and the integration of proton radiation [10] have not eliminated the risk of OM, underscoring the need for effective preventive interventions.

Current OM management strategies are largely supportive, including topical rinses, mouthwashes, and/or systemic narcotic analgesics, and are primarily aimed at symptom relief than preventing or treating OM [11]. Benzydamine mouthwash has been suggested for OM prevention; however, recommendations are limited to patients receiving moderate RT doses (<50 Gy) [12] and additional studies evaluating its efficacy have been recommended [13]. Keratinocyte growth factor was studied in head and neck cancer patients receiving definitive or post-op chemoradiation; however, it has not gained widespread acceptance due to its systemic administration requirements and risk-profile [14], [15]. Consequently, there remains a significant unmet need for an intervention that can be delivered reliably and safely to prevent OM in patients undergoing head and neck RT.

Clinical practice guidelines from the Multinational Association of Supportive Care in Cancer and the International Society of Oral Oncology (MASCC/ISOO) recommend intraoral photobiomodulation (PBM) therapy for the prevention of OM in patients undergoing head and neck RT, with or without concomitant chemotherapy, when delivered using specific treatment parameters [12]. These recommendations, subsequently adopted by the World Association for Photobiomodulation Therapy (WALT) [16], are supported by a substantial body of clinical evidence demonstrating that PBM therapy reduces the incidence and severity of OM, decreases pain and dysphagia, and improves quality of life in patients receiving head and neck RT [17], [18], [19], [20], [21].

PBM therapy uses non-ionizing red or near-infrared light to modulate inflammation, reduce pain and promote tissue repair [22], [23], [24], [25], [26]. However, despite strong efficacy data and guideline endorsement, PBM implementation in routine clinical practice remains limited, in part due to clinical workflow burden associated with daily treatment, challenges in treating the entire oral cavity, and variability in treatment administration across clinical settings.

An intraoral LED-based PBM system was developed to deliver PBM therapy to the oral cavity in a standardized manner prior to daily RT. This article reports the results of a double-blind, randomized, sham-controlled clinical trial evaluating the safety and effectiveness of the intraoral LED-based PBM device for reducing the incidence and severity of RT-induced OM and associated complications in patients with HNC.

## Methods

This prospective, multicenter, randomized (1:1), double-blind, sham-controlled clinical trial (NCT03972527) was conducted at 12 US centers, following Ethical Committee approval from the WIRB-Copernicus Group Institutional Review Board, NYU Langone Health, and MetroHealth. All participants provided written informed consent.

### Participants

Major inclusion criteria included patients diagnosed with squamous cell carcinoma of the oral cavity or oropharynx and planned to receive high dose intensity-modulated radiation therapy (IMRT) over 6-8-weeks, with at least two oral regions receiving >50 Gy, with or without concurrent chemotherapy.

Major exclusion criteria included prior RT to the head and neck, neoadjuvant or induction chemotherapy prior to starting RT, intensity modulated proton therapy, the presence of OM at baseline, smoking during IMRT, and medications indicated for the treatment and/or prevention of mucositis.

### Randomization and Blinding

Participants were randomized 1:1 to either the active photobiomodulation (PBM) therapy arm or control (sham) arm. Randomization was stratified according to tumor location and concurrent chemotherapy. Participants and investigators were blinded to the treatment allocation; only treatment administrators were unblinded. All participants were required to wear opaque goggles that obscured their vision and emitted a low-intensity red light to mask the treatment allocation.

### Study Interventions

Participants in both arms received a 10-minute study treatment session prior to each RT session, from the first to the last day of RT. For the active PBM arm, treatment was delivered using two mouthpieces containing three silicone-based optical waveguides that distributed light to target oral cavity tissues (**Figure 1**). The mouthpieces were used sequentially, and light was delivered at a wavelength of 660 nm over a 5-minute cycle to provide a nominal dose of 6 J/cm^2^. For the control arm, the mouthpieces were identical in appearance and utilization but modified for no light emission. The mouthpieces were reused for the same patient and underwent high-level disinfection between uses.

**Figure 1.**
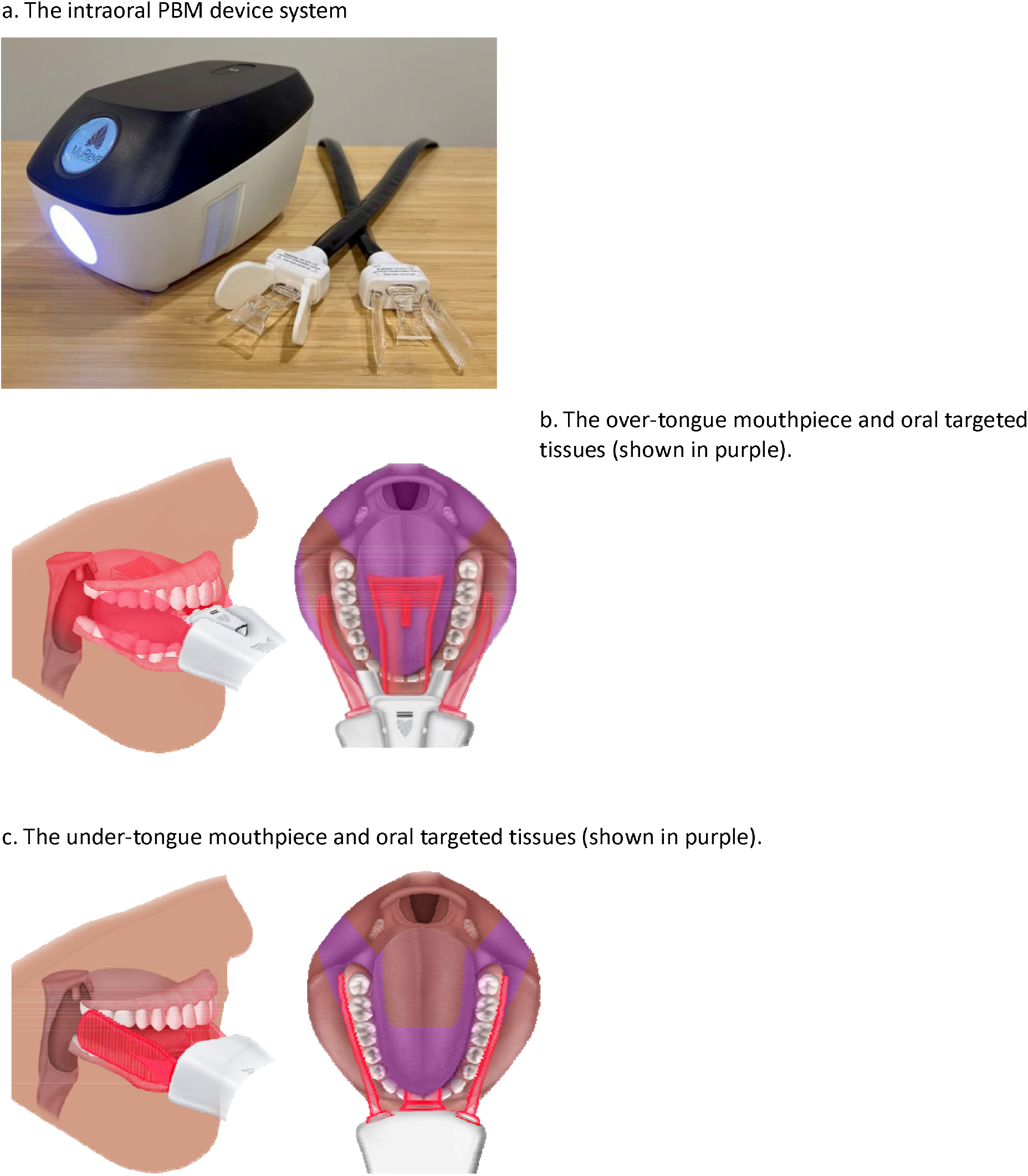
The intraoral PBM device system and OM-susceptible oral tissues targeted with system. (a) Picture of intraoral PBM device system (accessories not shown). The mouthpieces contain three silicone-based optical waveguides that distribute light to target tissues in the oral cavity. (b) The over-tongue mouthpiece targets the buccal mucosa, tongue, hard and soft palate, and oropharynx. (c) The under-tongue mouthpiece targets the ventral and lateral tongue, floor of mouth, and submandibular glands.

All participants followed standard-of-care oral hygiene and oral pain management protocols. A follow-up assessment was conducted two weeks after the completion of radiation therapy, consistent with standard post-treatment evaluation schedules.

### Outcome Assessments

#### Oral Mucositis

Oral mucositis was assessed at baseline, weekly during RT, and two weeks post-RT using the World Health Organization (WHO) Oral Toxicity Scale [27] and the Oral Mucositis Index [28]. All investigators underwent standardized training to ensure consistent scoring. At each assessment, investigators confirmed whether dietary limitations, if present, were attributable to oral complications.

The study included a pre-specified subgroup analysis of the primary endpoint stratified by radiation field size. The oral cavity was divided into nine anatomical regions corresponding to those assessed by the Oral Mucositis Index (upper labial mucosa, lower labial mucosa, right and left buccal mucosa, dorsal, lateral, and ventral tongue, floor of mouth, and soft palate). Participants were categorized into two groups (< median vs. ≥ median) based on the number of oral cavity regions expected to receive at least 30 Gy, as specified in the radiation treatment plan.

#### Patient-Reported Outcomes

Patient-reported assessments were performed at baseline, weekly during RT, and two weeks post-RT. Validated instruments were used to evaluate quality-of-life metrics. Mouth and throat soreness, throat pain, and OM symptom burden were assessed using the Oral Mucositis Weekly Questionnaire for Head and Neck Cancer (OMWQ-HN) [29]. Taste changes were assessed using the University of Washington Quality of Life Questionnaire (UoW QoL v4.1) [30].

Mouth and throat soreness (OMWQ-HN Question 3) was rated on a 5-point scale (0 = no soreness, 4 = extreme soreness). Throat pain (OMWQ-HN Question 6) was rated on an 11-point scale (0 = no pain or soreness, 10 = worst imaginable pain or soreness). Overall OM symptom burden was calculated by summing all OMWQ-HN item scores, excluding oral health, overall QOL, and limits on brushing teeth, using the method validated by Epstein (score range, 0-54) [29]. Taste ability (UoW QOL v4.1 Question 9) was rated as “I can taste food normally,”, “I can taste most foods normally,”, “I can taste some foods,” or “I cannot taste any foods.”

#### Safety

Adverse events were comprehensively recorded from the date of obtaining informed consent through study completion. Device safety was assessed through anticipated device-related Adverse Events (AEs) and Unanticipated Adverse Device Effects (UADEs). All device-related AEs and UADEs were assessed using the National Cancer Institute Common Terminology Criteria for Adverse Events (NCI CTCAE) v5.0. Adverse events were assessed by blinded investigators and independently reviewed by the blinded sponsor’s physicians.

### Sample Size and Statistical Analysis

A total of 82 participants were planned for enrollment. The sample size was determined based on the primary effectiveness endpoint of the mean OMI scores after six weeks of RT. Assuming a 7-point difference between the treatment and control arms, enrollment of 82 participants randomized 1:1 was estimated to provide a minimum statistical power of 91.3% to detect a statistically significant difference at a two-sided α = 0.05.

Three analysis populations were defined. The intent-to-treat (ITT) population included all enrolled and randomized participants. The per-protocol (PP) population included all enrolled and randomized participants who were compliant with the study procedures and had no major protocol deviations. The safety population included all enrolled and randomized participants who initiated at least one treatment session.

For the ITT population, missing values were imputed using a multiple imputation procedure utilizing predictive mean matching and either 50 or 100 sets of multiple imputations were generated depending on the analysis. An analysis was conducted for each set, and a pooled estimate of the test statistics was computed, along with a 95% interval estimate and p-value. Arm comparisons used Welch’s two-sample *t*-test, Mann-Whitney test, Fisher’s Exact Test, or *z*-test as appropriate. Statistical significance was defined as *p* < 0.05. All statistical analyses were performed using R, version 4.1.1 (R Foundation for Statistical Computing, Vienna, Austria).

## Results

Between August 2022 and July 2024, 100 patients were pre-screened for study eligibility, and a total of 85 patients provided written informed consent (**Table 1**) and were randomized to either the treatment arm (*n* = 42) or the control arm (*n* = 43) (**Fig. 2**). The study experienced 10 subject discontinuations, with 3 in the treatment arm and 7 in the control arm. Reasons for withdrawal varied and there were no consistent trends observed in the reasons for subject withdrawal. All withdrawn subjects were part of the ITT analysis.

**Table 1.** Patient and Demographic Characteristics by Study Arm for Intent-to-Treat Population.

|  | Treatment Arm (n=42)<br>n (%) | Control Arm (n=43)<br>n (%) | p - value |
| --- | --- | --- | --- |
| Age, mean ± s.d. (range) | 59.7 ± 13.4 (24-87) | 59.3 ± 10.7 (25-78) | |
| < 30 | 3 (7.1) | 1 (2.3) |  |
| 31-40 | 1 (2.4) | 1 (2.3) |  |
| 41-50 | 2 (4.8) | 7 (16.3) | 0.281 |
| 51-60 | 12 (28.6) | 15 (34.9) |  |
| 61-70 | 15 (35.7) | 13 (30.2) |  |
| 71-80 | 8 (19.0) | 6 (14.0) |  |
| > 80 | 1 (2.4) | 0 (0.0) |  |
| Sex |  |  |  |
| Female | 12 (28.6) | 12 (27.9) | 1 |
| Male | 30 (71.4) | 31 (72.1) |  |
| Race |  |  |  |
| African American | 3 (7.1) | 2 (4.7) |  |
| American Indian | 0 (0.0) | 1 (2.3) | 0.458 |
| Caucasian | 35 (83.3) | 38 (88.4) |  |
| Mixed | 0 (0.0) | 1 (2.3) |  |
| Other | 4 (9.5) | 1 (2.3) |  |
| Ethnicity |  |  |  |
| Hispanic or Latino | 4 (9.5) | 4 (9.3) | 1 |
| Not Hispanic or Latino | 38 (90.5) | 39 (90.7) |  |
| Prior Nicotine Use |  |  |  |
| Quit >2 years from treatment start | 10 (47.6) | 14 (45.2) | 1 |
| Quit <2 years from treatment start | 11 (52.4) | 17 (54.8) |  |
| T Stage |  |  |  |
| 0 | 1 (2.4) | 2 (4.7) |  |
| 1 | 6 (14.3) | 5 (11.6) | 0.462 |
| 2 | 20 (47.6) | 13 (30.2) |  |
| 3 | 9 (21.4) | 15 (34.9) |  |
| 4 | 6 (14.3) | 8 (18.6) |  |
| Primary Tumor Location |  |  |  |
| Oral Cavity | 12 (28.6) | 10 (23.3) | 0.860 |
| Non-HPV related Oropharyngeal Cancer | 5 (11.9) | 6 (14.0) |  |
| HPV-related Oropharyngeal Cancer | 25 (59.5) | 27 (62.8) |  |
| HPV Status |  |  |  |
| No | 11 (26.2) | 9 (20.9) | 0.856 |
| Yes | 26 (61.9) | 28 (65.1) |  |
| Unknown | 5 (11.9) | 6 (14.0) |  |
| AJCC Stage |  |  |  |
| Early-Stage (I – II) | 7 (16.7) | 5 (11.6) | 0.549 |
| Advanced-Stage (III – IVb) | 35 (83.3) | 38 (88.4) |  |
| Treatment Type |  |  |  |
| Adjuvant | 13 (31.0) | 12 (27.9) | 0.815 |
| Definitive | 29 (69.0) | 31 (72.1) |  |
| Number of oral sites receiving >30 Gy of radiation |  |  |  |
| < Median | 21 (50.0) | 19 (44.2) | 0.666 |
| ≥ Median | 21 (50.0) | 24 (55.8) | |
| Number of oral sites receiving >50 Gy of radiation |  |  |  |
| 2 | 9 (21.4) | 8 (18.6) |  |
| 3 | 15 (35.7) | 18 (41.9) | 0.610 |
| 4 | 9 (21.4) | 12 (27.9) |  |
| 5 | 9 (21.4) | 5 (11.6) |  |
| Chemotherapy use during cancer treatment |  |  |  |
| No | 9 (21.4) | 10 (23.3) | 1 |
| Yes | 33 (78.6) | 33 (76.7) |  |
| Radiation exposure after 6 weeks of RT (Gy), mean ± s.d. (median) | 56.8 ± 4.6 (57.2) | 57.1 ± 4.0 (57.6) | 0.856 |
| Total radiation exposure (Gy), mean ± s.d. (median) | 66.8 ± 4.6 (70.0) | 66.7 ± 4.5 (70.0) | 0.934 |
| Total days of RT, mean ± s.d. (median) | 33.3 ± 2.0 (33.0) | 33.0 ± 2.1 (33.0) | 0.580 |

**Figure 2.**
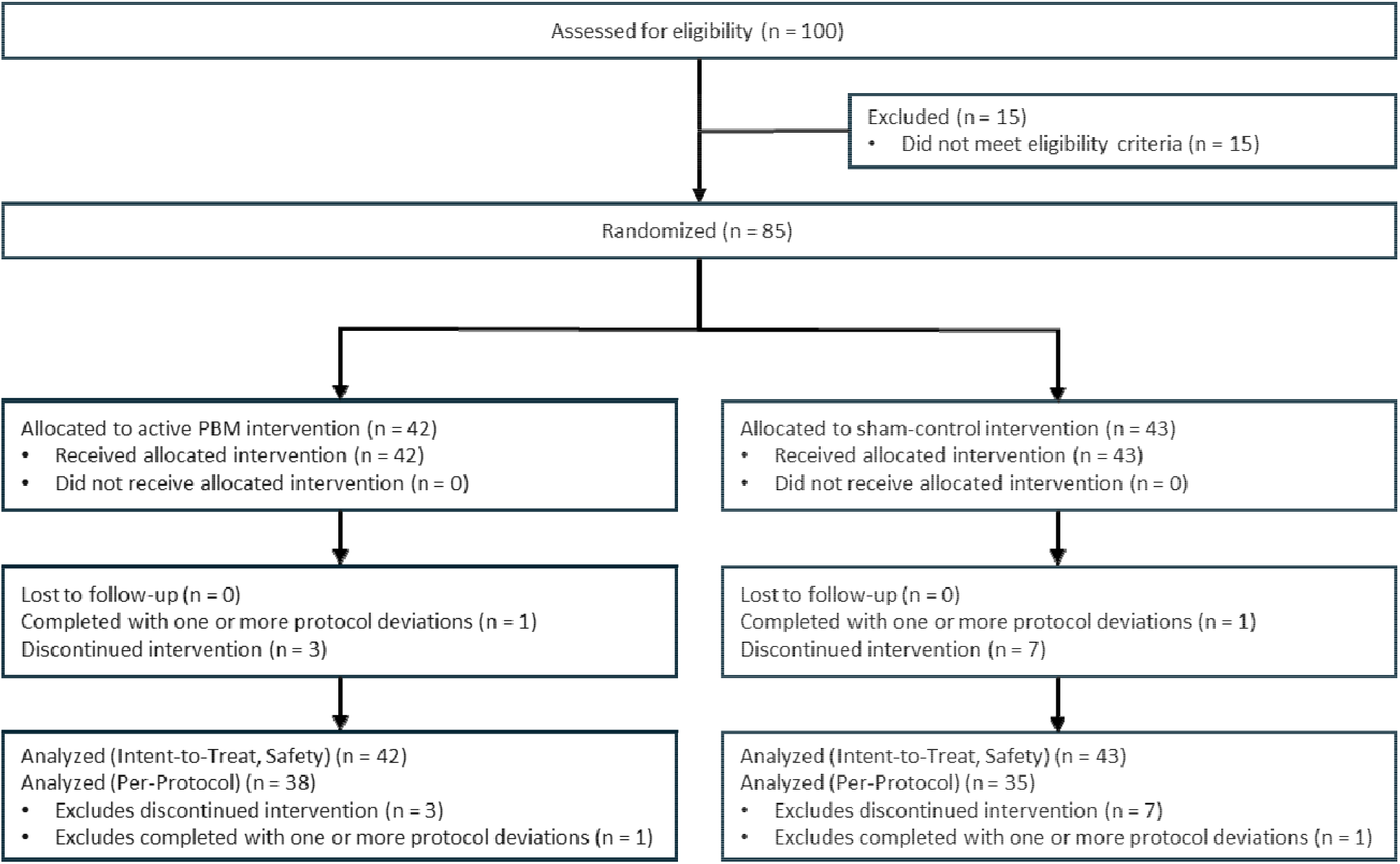
CONSORT Diagram.

No significant differences in demographics or in disease- or treatment-related factors were observed between the two arms. The total radiation dose delivered, dose after six weeks, number of RT treatment days, and number of oral sites receiving >30 or >50 Gy of radiation were similar between the two arms.

### Efficacy Outcomes

#### Primary Endpoint – Oral Mucositis Index Scores After Six Weeks of IMRT

All participants had an OMI score of 0 at baseline. While participants randomized to the treatment arm exhibited lower mean OMI scores at six weeks of treatment, this did not reach statistical significance (**Table 2**). As part of the primary endpoint analysis, a pre-specified subgroup analysis was conducted to evaluate the outcomes based on the extent of oral cavity radiation exposure. Among the participants who received subtotal oral cavity irradiation (>30 Gy to <75% of the oral cavity), treatment arm participants had significantly lower OMI scores at six weeks than the control arm (10.2 in treatment arm vs. 15.3 in control arm; ITT, *p* = 0.023) (**Table 2**).

**Table 2.** Oral Mucositis Index (OMI) Scores after 6 Weeks of Treatment. The study included a pre-specified subgroup analysis of the primary endpoint stratified by radiation field size. The oral cavity was divided into nine anatomical regions corresponding to those assessed by the OMI (upper labial mucosa, lower labial mucosa, right and left buccal mucosa, dorsal, lateral, and ventral tongue, floor of mouth, and soft palate). Participants were categorized into two groups (< median vs. ≥ median) based on the number of oral cavity regions expected to receive at least 30 Gy, as specified in the radiation treatment plan.

| Subject Population | Oral Mucositis Index (OMI) score after 6 weeks of treatment |  |  |  |  |  | *p* - value |  |
| --- | --- | --- | --- | --- | --- | --- | --- | --- |
|  | Treatment Arm* |  |  | Control Arm* |  |  |  |  |
|  | n | Mean | SD | n | Mean | SD | ITT | PP |
| All randomized participants | 39 | 12.3 | 7.9 | 36 | 14.0 | 7.0 | 0.338 | 0.346 |
| Participants receiving >30 Gy of radiation to < 75% of oral cavity | 20 | 10.2 | 6.0 | 16 | 15.3 | 6.1 | 0.023 | 0.014 |
| Participants receiving >30 Gy of radiation to > 75% of oral cavity | 19 | 14.6 | 9.1 | 20 | 13.0 | 7.7 | 0.420 | 0.549 |

\* N, mean, and SD values are shown for study completers. Missing values in the ITT population were imputed using predictive mean matching with 50 datasets per analysis.

#### Severe Oral Mucositis Incidence During Six Weeks of IMRT (WHO Oral Toxicity Scale)

A reduction in the incidence of severe oral mucositis was observed in the treatment arm compared with that in the control arm. Across the first six weeks of IMRT, 36.8% of participants in the treatment arm and 57.1% in the control arm developed severe OM as assessed using the WHO Oral Toxicity Scale (ITT, *p* = 0.046) (**Figure 3A**). Weekly analyses of the per-protocol (PP) population demonstrated lower severe OM rates in the treatment arm during weeks 3–6 of IMRT (**Supplemental Figure 1**). None of the participants in either arm developed severe OM during the first two weeks of treatment.

**Figure 3.**
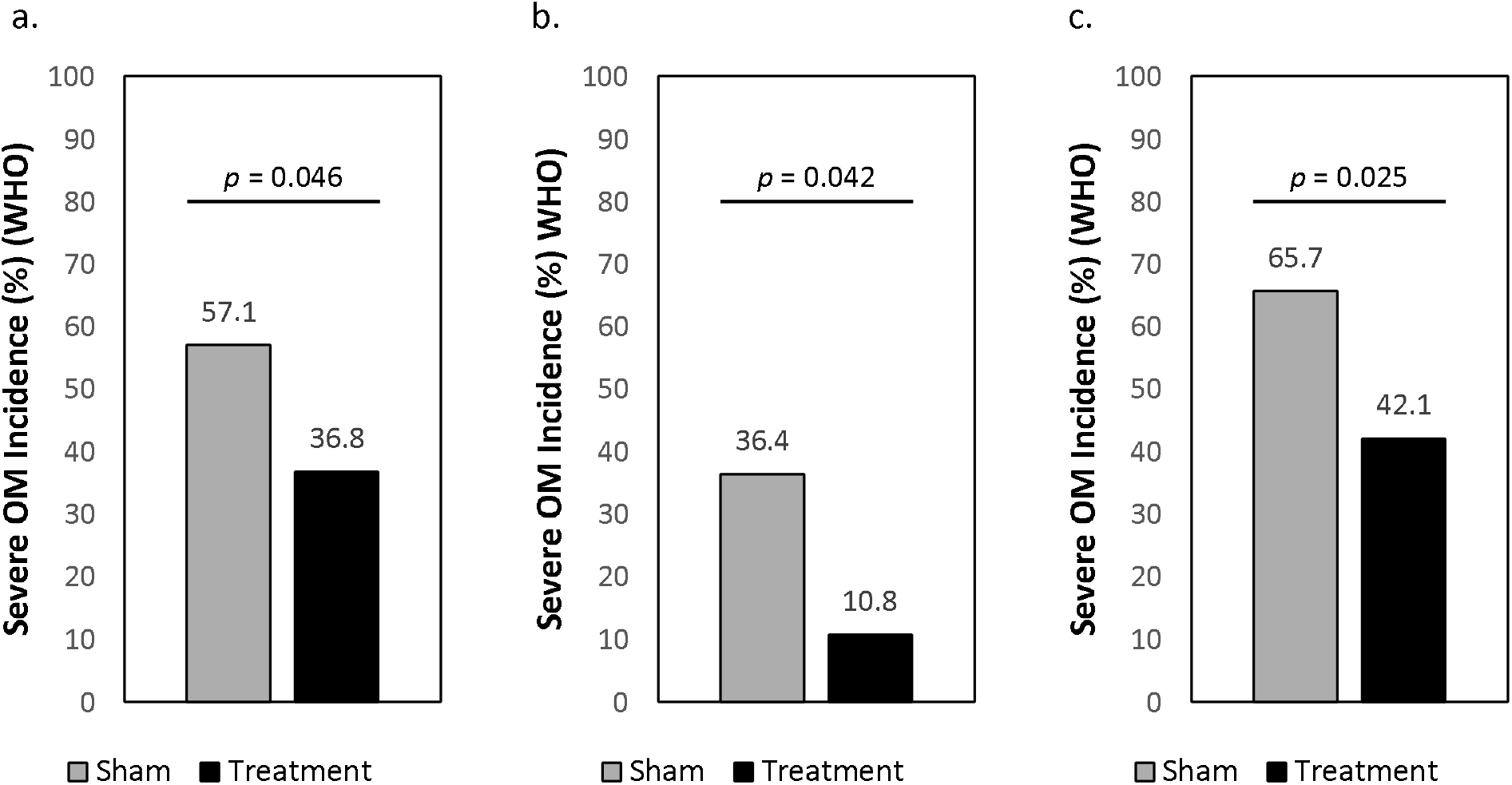
Difference in severe oral mucositis (OM) incidence as assessed using the WHO Oral Toxicity (WHO) Scale (ITT population). (a) Severe OM Incidence during six weeks of intensity-modulated radiation therapy. (b) Severe OM incidence at two-week post-radiation therapy (RT) visit. (c) Severe OM incidence from baseline through two-weeks post-RT.

#### Severe Oral Mucositis Incidence Two Weeks Following Cancer Treatment (WHO Oral Toxicity Scale)

Two weeks after cancer treatment concluded, the incidence of severe OM was 10.8% in the treatment arm and 36.4% in the control arm (ITT, *p* = 0.042) (**Figure 3B**).

#### Severe Oral Mucositis Incidence During Cancer Treatment Through Two-Week Post-RT Visit (WHO Oral Toxicity Scale)

For participants whose cancer treatment extended beyond six weeks, a separate analysis was conducted to encompass the entire duration of study participation, including both the active cancer treatment phase and the two-week follow-up period. The incidence of severe mucositis was reduced from 65.7% in the control arm to 42.1% in the treatment arm (ITT, *p* = 0.025) (**Figure 3C**).

#### Duration of Severe Oral Mucositis (WHO Oral Toxicity Scale)

The duration of severe OM was evaluated from treatment initiation through two weeks post-RT. Participants in the treatment arm experienced severe OM for a median of 16.2% of their study participation duration, compared with 28.3% in the control arm (ITT, *p* = 0.060; PP, *p* = 0.043) (**Supplemental Figure 2**).

#### Changes in Patient-Reported Outcomes After 6 Weeks of IMRT

Over the six-week course of RT, participants in the treatment arm reported a 32% smaller mean increase in soreness over six weeks than those in the control arm (1.5 vs. 2.2; PP, *p* = 0.029) (**Figure 4A**). Changes in throat pain followed a similar pattern. The participants in the treatment arm reported a 28% smaller mean increase in throat pain than those in the control arm (3.8 vs. 5.3; PP, *p* = 0.028) (**Figure 4B**).

**Figure 4.**
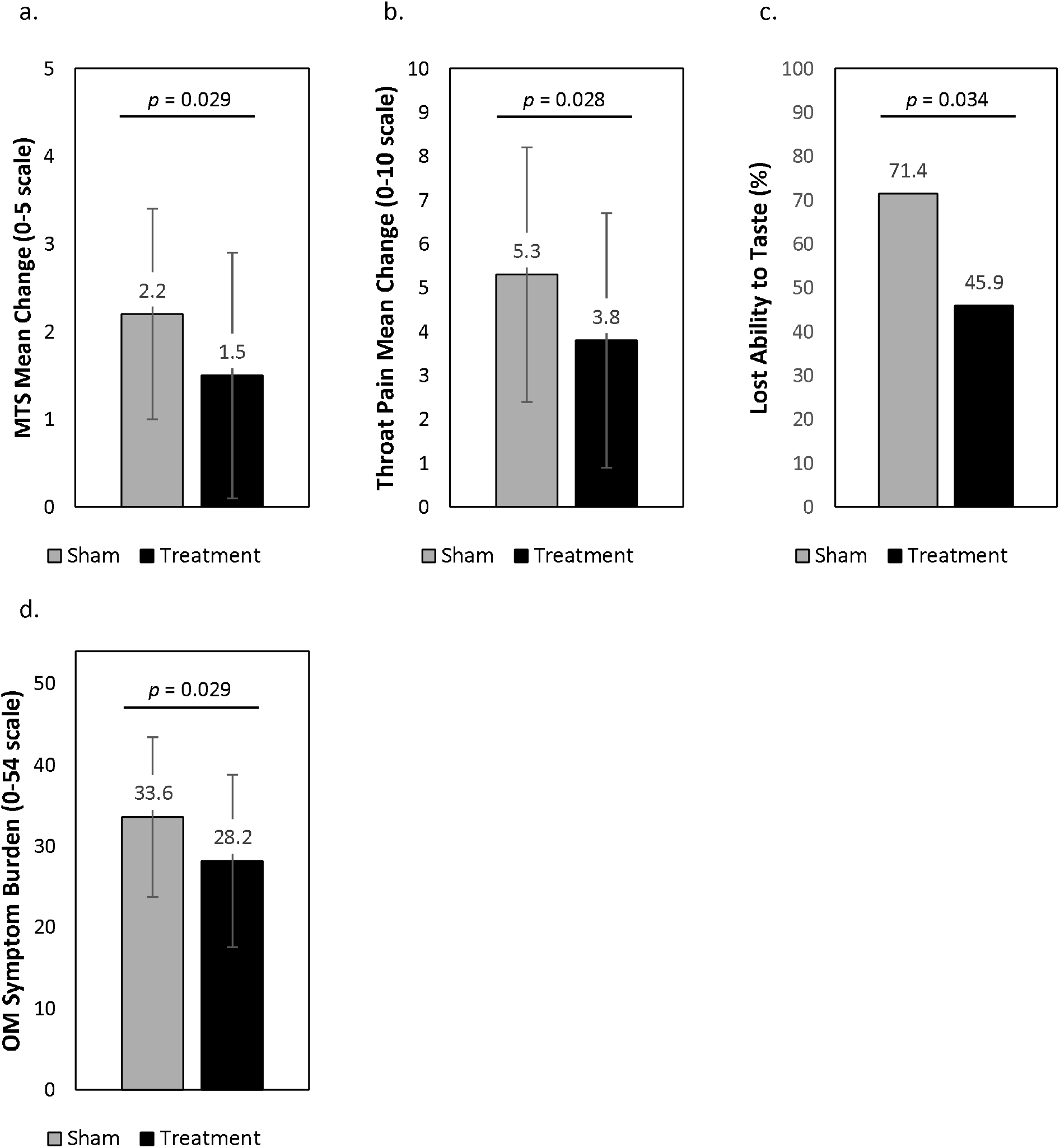
Patient-reported outcomes after six weeks of intensity-modulated radiation therapy (PP Population). (a) Changes in Mouth and Throat Soreness (MTS) from baseline (OMWQ-HN Question 3). (b) Changes in Throat Pain from baseline (OMWQ-HN Question 6). (c) Loss of taste (UoW QoL v4.1 Question 9). (d) Symptom burden (OMWQ-HN).

After six weeks of cancer treatment, participants randomized to the treatment arm exhibited a 35.7% reduction in taste loss relative to the control arm (45.9% vs. 71.4%; PP, *p* = 0.034) (**Figure 4C**). Further, OM symptom burden was reduced 16.1% (28.2 ± 10.7 vs. 33.7 ± 10.0; PP, *p* = 0.037) (**Figure 4D**).

#### Reduction in Medical Interventions

Surgical feeding tube placement was recorded from the date of consent through the final two weeks post-RT visit. 21.2% of participants (18 of 85) had a feeding tube placed prior to the start of IMRT. After the start of RT, there was a 59% reduction in surgical feeding tube placement in the treatment arm compared with the control arm (15.2% vs. 37%; PP, *p* = 0.073), representing a strong trend albeit not reaching statistical significance (**Supplemental Figure 3**).

### Treatment Adherence and Tolerability

The device was well tolerated by all participants (treatment or control). Of the 2,574 device sessions initiated, 99.5% (2,561/2,574) were completed. No difference in completion rates was observed between the study arms (99.5% treatment vs. 99.5% control, *p* = 1, Fisher’s exact test). No procedural modifications were required for participants with anatomical variations (e.g., tongue tie, post-surgical resection, and edentulism) (**Supplemental Table 1**).

### Safety Evaluation

Treatment was safe and there were no device-related adverse events.

## Discussion

In this randomized, double-blind, sham-controlled trial, intraoral LED-based PBM therapy was associated with a reduction in the incidence of severe OM and OM-related complications in patients with HNC undergoing IMRT, with or without concurrent chemotherapy. The intervention was well tolerated, with no device-related adverse events observed. Given the frequency and clinical impact of OM during head and neck RT, these findings provide evidence that PBM therapy can reduce treatment-related morbidity in this population.

Patients receiving daily PBM therapy experienced a significantly lower incidence of severe OM (WHO Grade ≥3) from IMRT initiation through two weeks post-RT compared with the control arm. The incidence of severe OM was reduced from 65.7% in the control arm to 42.1% in the PBM arm. Further, severe OM incidence two weeks post-RT was reduced by 70% in the PBM arm. These findings were accompanied by favorable outcomes in objective mucosal injury scores, reduced severe OM duration, patient-reported outcomes, and a reduced need for additional medical interventions to support nutrition. Among patients receiving subtotal oral cavity irradiation (>30 Gy to <75% of the oral cavity), mean OMI scores at six weeks of RT were 33% lower in the treatment arm (ITT, *p* = 0.023), suggesting a measurable reduction in mucosal ulceration and erythema.

Patient-reported outcomes after six weeks of IMRT further support the clinical relevance of these findings. Participants receiving PBM therapy reported significantly smaller increases from baseline in mouth and throat soreness and throat pain compared with the control arm. Cumulatively, there was a significant reduction in OM symptom burden in patients receiving PBM therapy. Notably, this trial is among the first to demonstrate greater preservation of taste in patients receiving PBM therapy, an outcome with important implications for maintenance of nutritional status and quality of life.

A 59% reduction in surgical feeding tube placement after the start of RT was also observed in the PBM arm; however, despite the magnitude of this difference (15.2% in the PBM arm vs. 37% in the control arm), statistical significance was not achieved. While this outcome should be interpreted cautiously, the observed preservation of nutritional intake without escalation to invasive interventions warrants further investigation, as it may suggest a potential association between daily PBM therapy and reduced need for feeding tube placement in selected patients. This observation is further supported by the concurrent reductions in severe OM and favorable patient-reported outcomes observed in the PBM arm. Future analyses should assess outcomes separately among patients initiating RT without a feeding tube.

No OM-related treatment interruptions were observed in either arm. The absence of interruptions in the control arm was unexpected given the high incidence of severe OM and prior reports linking severe OM to treatment modification or dose interruption [31], [32]. This finding warrants further investigation, particularly given evidence that unplanned treatment breaks are associated with poorer clinical outcomes [33]. Collectively, these findings support intraoral PBM as an effective supportive care strategy to reduce OM severity, preserve oral function, and decrease escalation of supportive interventions during head and neck RT.

The results of this study are consistent with the broader literature supporting the efficacy of PBM therapy for the prevention and treatment of cancer therapy-induced OM. This literature formed the basis for current MASCC/ISOO and WALT guideline recommendations for PBM in patients undergoing head and neck RT, with or without concurrent chemotherapy. These guidelines provide specific technical parameters for use of handheld probes including wavelength, power density, spot size, energy density, number of oral sites treated, and distance from the tissue. Variability in any of these technical parameters can adversely influence therapeutic outcomes, necessitating trained operators and contributing to challenges in reproducibility. As most protocols for patients with HNC involve PBM delivered 5 days per week for the entire 6-8-week course of RT, the administration of PBM to these patients using handheld probes is highly labor-intensive, resource-intensive, and logistically challenging. As a result, PBM therapy has not been widely adopted in routine clinical practice despite strong clinical support.

In this study, treatment with the intraoral PBM device was feasible within a standard daily radiation therapy workflow and was well tolerated by participants. Treatment sessions were completed as scheduled using a standardized, dose-controlled 10-minute protocol targeting all OM-susceptible tissues, without the need for procedural modifications across a range of oral anatomies. These findings suggest that intraoral PBM therapy can be implemented consistently in both academic and community-based clinical settings.

The study has several strengths, including its randomized, double-blind, sham-controlled design, which significantly reduced bias in the study assessments. Randomization and stratification resulted in balanced treatment arms with respect to known OM risk factors. Reductions in objective measures of OM severity were concordant with improvements in patient-reported outcomes, supporting the internal consistency of the findings. A limitation of this study is that the primary endpoint of mean OMI scores at six weeks of RT did not achieve statistical significance for the overall population. However, mean OMI scores were significantly lower among patients receiving subtotal oral cavity irradiation (>30 Gy to <75% of the oral cavity), while variability was high among those receiving total irradiation (>30 Gy to >75% of the oral cavity), particularly in the treatment arm. In this subgroup, a greater proportion of treatment arm participants had high-dose irradiation (>60 Gy) to the dorsal, ventral, and lateral tongue, regions that account for 40% of the OMI score. The combined influence of anatomic weighting and dose intensity may have contributed to the observed variability. In addition, larger radiation fields may be associated with a smaller pool of recruitable healthy cells, potentially attenuating the tissue repair response associated with PBM therapy. These findings indicate that a larger sample size may be needed to more fully characterize treatment effects in patients undergoing total oral cavity irradiation.

In summary, systematic reviews of randomized controlled trials have concluded that PBM therapy is beneficial for the prevention and/or treatment of OM [6], [34], [35]. This randomized, double-blind, sham-controlled trial provides evidence supporting the safety and effectiveness of intraoral PBM therapy for reducing the incidence and severity of radiation-induced OM in patients with HNC. These findings align with current clinical practice guidelines and support the integration of PBM into evidence-based supportive care strategies for patients undergoing radiation therapy.

## Data Availability

The data that supports the findings reported in this paper are not publicly available because they contain information that could compromise the privacy of the study participants. Data are available from the corresponding author (K.H.) upon reasonable request.

## Acknowledgements

We are grateful to all the patients who participated in the present trial and to the study personnel for their support. Statistical analyses were performed by Peter Kupchak, PhD (Guelph, Ontario, Canada). Medical writing assistance was provided by Larry Yost from The Atticus Group, LLC (Portsmouth, New Hampshire, USA).

## References

[1] L. S. Elting, C. D. Cooksley, M. S. Chambers, and A. S. Garden, “Risk, Outcomes, and Costs of Radiation-Induced Oral Mucositis Among Patients With Head-and-Neck Malignancies,” International Journal of Radiation Oncology*Biology*Physics, vol. 68, no. 4, pp. 1110–1120, Jul. 2007, doi: 10.1016/j.ijrobp.2007.01.053.

[2] B. A. Murphy, “Clinical and economic consequences of mucositis induced by chemotherapy and/or radiation therapy.,” J. Support. Oncol., vol. 5, no. 9 Suppl 4, pp. 13–21, Oct. 2007.

[3] M. U. R. Naidu, G. V. Ramana, P. U. Rani, lyyapu K. Mohan, A. Suman, and P. Roy, “Chemotherapy-Induced and/or Radiation Therapy-Induced Oral Mucositis-Complicating the Treatment of Cancer,” Neoplasia, vol. 6, no. 5, pp. 423–431, Sep. 2004, doi: 10.1593/neo.04169.

[4] A. F. Lopes Martins et al., “Cost-effectiveness randomized clinical trial on the effect of photobiomodulation therapy for prevention of radiotherapy-induced severe oral mucositis in a Brazilian cancer hospital setting,” Supportive Care in Cancer, vol. 29, no. 3, pp. 1245–1256, Mar. 2021, doi: 10.1007/s00520-020-05607-6.

[5] K. Berger et al., “Burden of Oral Mucositis: A Systematic Review and Implications for Future Research,” Oncol. Res. Treat., vol. 41, no. 6, pp. 399–405, 2018, doi: 10.1159/000487085.

[6] S. Parra-Rojas, J. Cassol Spanemberg, N. del Mar Díaz-Robayna, M. Peralta-Mamani, and R.T. Velázquez Cayón, “Assessing the Cost-Effectiveness of Photobiomodulation for Oral Mucositis Prevention and Treatment: A Systematic Review,” Biomedicines, vol. 12, no. 10, p. 2366, Oct. 2024, doi: 10.3390/biomedicines12102366.

[7] L. Rodrigues-Oliveira et al., “Direct costs associated with the management of mucositis: A systematic review,” Oral Oncol., vol. 118, p. 105296, Jul. 2021, doi: 10.1016/j.oraloncology.2021.105296.

[8] S. S. Yom et al., “Reduced-Dose Radiation Therapy for HPV-Associated Oropharyngeal Carcinoma (NRG Oncology HN002),” Journal of Clinical Oncology, vol. 39, no. 9, pp. 956–965, Mar. 2021, doi: 10.1200/JCO.20.03128.

[9] R. L. Ferris et al., “Phase II Randomized Trial of Transoral Surgery and Low-Dose Intensity Modulated Radiation Therapy in Resectable p16+ Locally Advanced Oropharynx Cancer: An ECOG-ACRIN Cancer Research Group Trial (E3311),” Journal of Clinical Oncology, vol. 40, no. 2, pp. 138– 149, Jan. 2022, doi: 10.1200/JCO.21.01752.

[10] S. J. Frank et al., “Proton versus photon radiotherapy for patients with oropharyngeal cancer in the USA: a multicentre, randomised, open-label, non-inferiority phase 3 trial,” The Lancet, Dec. 2025, doi: 10.1016/S0140-6736(25)01962-2.

[11] M. Cinausero et al., “New Frontiers in the Pathobiology and Treatment of Cancer Regimen-Related Mucosal Injury,” Front. Pharmacol., vol. 8, Jun. 2017, doi: 10.3389/fphar.2017.00354.

[12] S. Elad et al., “MASCC/ISOO clinical practice guidelines for the management of mucositis secondary to cancer therapy,” Cancer, vol. 126, no. 19, pp. 4423–4431, Oct. 2020, doi: 10.1002/cncr.33100.

[13] O. Nicolatou-Galitis, P. Bossi, E. Orlandi, and René-Jean Bensadoun, “The role of benzydamine in prevention and treatment of chemoradiotherapy-induced mucositis,” Supportive Care in Cancer, vol. 29, no. 10, pp. 5701–5709, Oct. 2021, doi: 10.1007/s00520-021-06048-5.

[14] M. Henke et al., “Palifermin Decreases Severe Oral Mucositis of Patients Undergoing Postoperative Radiochemotherapy for Head and Neck Cancer: A Randomized, Placebo-Controlled Trial,” Journal of Clinical Oncology, vol. 29, no. 20, pp. 2815–2820, Jul. 2011, doi: 10.1200/JCO.2010.32.4103.

[15] Q.-T. Le et al., “Palifermin Reduces Severe Mucositis in Definitive Chemoradiotherapy of Locally Advanced Head and Neck Cancer: A Randomized, Placebo-Controlled Study,” Journal of Clinical Oncology, vol. 29, no. 20, pp. 2808–2814, Jul. 2011, doi: 10.1200/JCO.2010.32.4095.

[16] J. Robijns et al., “Photobiomodulation therapy in management of cancer therapy-induced side effects: WALT position paper 2022,” Front. Oncol., vol. 12, Aug. 2022, doi: 10.3389/fonc.2022.927685.

[17] H. T. Whelan et al., “Effect of NASA Light-Emitting Diode Irradiation on Wound Healing,” J. Clin. Laser Med. Surg., vol. 19, no. 6, pp. 305–314, Dec. 2001, doi: 10.1089/104454701753342758.

[18] T. Zanin et al., “Use of 660-nm Diode Laser in the Prevention and Treatment of Human Oral Mucositis Induced by Radiotherapy and Chemotherapy,” Photomed. Laser Surg., vol. 28, no. 2, pp. 233–237, Apr. 2010, doi: 10.1089/pho.2008.2242.

[19] P. A. G. Carvalho, G. C. Jaguar, A. C. Pellizzon, J. D. Prado, R. N. Lopes, and F. A. Alves, “Evaluation of low-level laser therapy in the prevention and treatment of radiation-induced mucositis: A double-blind randomized study in head and neck cancer patients,” Oral Oncol., vol. 47, no. 12, pp. 1176–1181, Dec. 2011, doi: 10.1016/j.oraloncology.2011.08.021.

[20] H. S. Antunes et al., “Phase III trial of low-level laser therapy to prevent oral mucositis in head and neck cancer patients treated with concurrent chemoradiation,” Radiotherapy and Oncology, vol. 109, no. 2, pp. 297–302, Nov. 2013, doi: 10.1016/j.radonc.2013.08.010.

[21] A. F. Oton-Leite, A.C. Corrêa de Castro, M. O. Morais, J. C. D. Pinezi, C. R. Leles, and E. F. Mendonça, “Effect of intraoral low-level laser therapy on quality of life of patients with head and neck cancer undergoing radiotherapy,” Head Neck, vol. 34, no. 3, pp. 398–404, Mar. 2012, doi: 10.1002/hed.21737.

[22] B. Joseph, M. Mauramo, T. Sorsa, S. Anil, and T. Waltimo, “LED-based low-level light therapy for oral mucositis in cancer patients: a systematic review and GRADE analysis,” Oral Surg. Oral Med. Oral Pathol. Oral Radiol., vol. 140, no. 3, pp. 300–311, Sep. 2025, doi: 10.1016/j.oooo.2025.04.095.

[23] S. Barati, S. Motevasseli, H. S. Saedi, P. Amiri, and R. Fekrazad, “Effectiveness of Photobiomodulation (low-level laser therapy) on treatment of oral mucositis (OM) induced by chemoradiotherapy in head and neck cancer patients.,” J. Photochem. Photobiol. B, vol. 264, p. 113115, Mar. 2025, doi: 10.1016/j.jphotobiol.2025.113115.

[24] R. Hanna et al., “Photobiomodulation Therapy in Oral Mucositis and Potentially Malignant Oral Lesions: A Therapy Towards the Future,” Cancers (Basel)., vol. 12, no. 7, p. 1949, Jul. 2020, doi: 10.3390/cancers12071949.

[25] E. Courtois et al., “Mechanisms of PhotoBioModulation (PBM) focused on oral mucositis prevention and treatment: a scoping review,” BMC Oral Health, vol. 21, no. 1, p. 220, Dec. 2021, doi: 10.1186/s12903-021-01574-4.

[26] B. Shen, Y. Zhou, D. Wu, and J. Liu, “Efficacy of photobiomodulation therapy in the management of oral mucositis in patients with head and neck cancer: A systematic review and meta-analysis of randomized controlled trials,” Head Neck, vol. 46, no. 4, pp. 936–950, Apr. 2024, doi: 10.1002/hed.27655.

[27] WHO handbook for reporting results of cancer treatment. World Health Organization[]; [Sold by WHO Publications Centre USA], 1979.

[28] A. Barasch et al., “Helium-neon laser effects on conditioning-induced oral mucositis in bone marrow transplantation patients,” Cancer, vol. 76, no. 12, pp. 2550–2556, Dec. 1995, doi: 10.1002/1097-0142(19951215)76:12<2550::AID-CNCR2820761222>3.0.CO;2-X.

[29] J. B. Epstein et al., “Longitudinal evaluation of the oral mucositis weekly questionnaire-head and neck cancer, a patient-reported outcomes questionnaire,” Cancer, vol. 109, no. 9, pp. 1914–1922, May 2007, doi: 10.1002/cncr.22620.

[30] S. N. Rogers, D. Lowe, B. Yueh, and E. A. Weymuller Jr, “The Physical Function and Social-Emotional Function Subscales of the University of Washington Quality of Life Questionnaire,” Arch. Otolaryngol. Head. Neck Surg., vol. 136, no. 4, p. 352, Apr. 2010, doi: 10.1001/archoto.2010.32.

[31] O. M. Maria, N. Eliopoulos, and T. Muanza, “Radiation-Induced Oral Mucositis,” Front. Oncol., vol. 7, May 2017, doi: 10.3389/fonc.2017.00089.

[32] T. Shaikh, E. A. Handorf, C. T. Murphy, R. Mehra, J. A. Ridge, and T. J. Galloway, “The Impact of Radiation Treatment Time on Survival in Patients With Head and Neck Cancer,” International Journal of Radiation Oncology*Biology*Physics, vol. 96, no. 5, pp. 967–975, Dec. 2016, doi: 10.1016/j.ijrobp.2016.08.046.

[33] L. A. Gharzai et al., “Treatment Interruption and Outcomes in Head and Neck Cancer,” JAMA Otolaryngology–Head & Neck Surgery, Dec. 2025, doi: 10.1001/jamaoto.2025.4203.

[34] Y. Zadik et al., “Systematic review of photobiomodulation for the management of oral mucositis in cancer patients and clinical practice guidelines,” Supportive Care in Cancer, vol. 27, no. 10, pp. 3969–3983, Oct. 2019, doi: 10.1007/s00520-019-04890-2.

[35] M. Gobbo et al., “Quality assessment of PBM protocols for oral complications in head and neck cancer patients: part 2,” Supportive Care in Cancer, vol. 31, no. 5, p. 306, May 2023, doi: 10.1007/s00520-023-07749-9.

