## Supplemental Materials for "Randomized, double-blind, sham-controlled trial of an intraoral photobiomodulation device for oral mucositis due to radiotherapy for head and neck cancer"

### Supplemental Table 1. Tolerability Results for Pre-Existing Dental / Oral Conditions.

| Pre-Existing Condition* | N | Potential Treatments | Treatments Declined | Declined % | Completed in Full | Not in Full | Completion % |
|---|---|---|---|---|---|---|---|
| Fully Edentulous | 18 | 520 | 5 | 1% | 511 | 4 | 99.2% |
| Tongue-Tied | 4 | 96 | 0 | 0% | 95 | 1 | 98.9% |
| Prior Resection | 13 | 364 | 2 | 0.5% | 361 | 1 | 99.7% |
| None | 53 | 1682 | 19 | 1.1% | 1654 | 9 | 99.5% |

*N, mean, and SD values are shown for study completers. Missing values in the ITT population were imputed using predictive mean matching with 50 datasets per analysis.

**Supplemental Figure 1.** Severe oral mucositis (OM) incidence by week as assessed using the WHO Oral Toxicity (WHO) Scale (PP population).

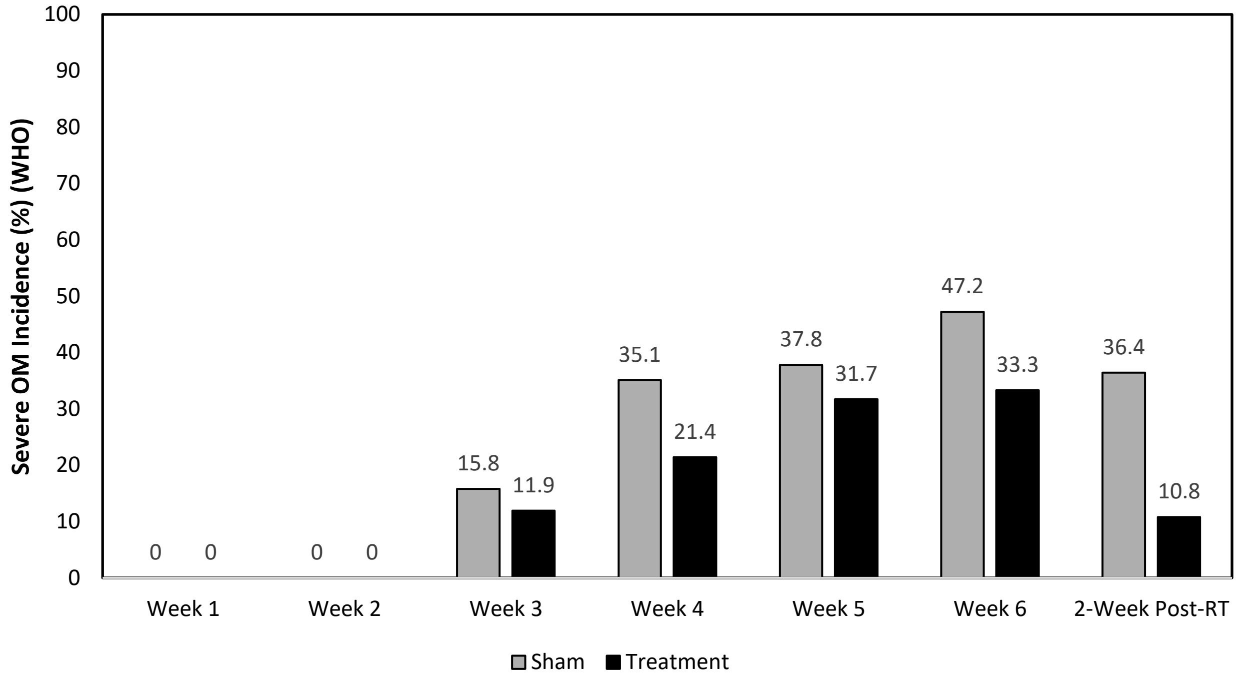

**Supplemental Figure 2.** Duration of severe oral mucositis from baseline through two-weeks post-radiation therapy as assessed using the WHO Oral Toxicity Scale (ITT population). Duration is represented as a percentage of the total period from baseline through two-weeks post-radiation therapy.

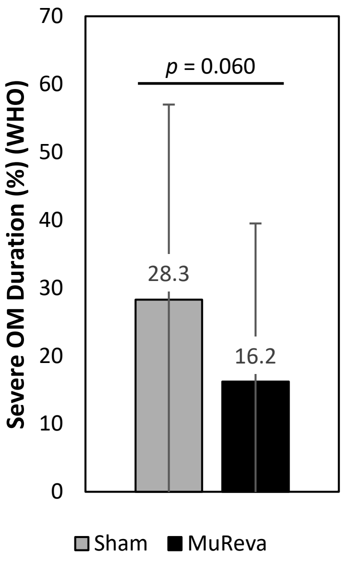

**Supplemental Figure 3.** Percentage of participants requiring surgical feeding tube placement during intensity-modulated radiation therapy* (PP Population).

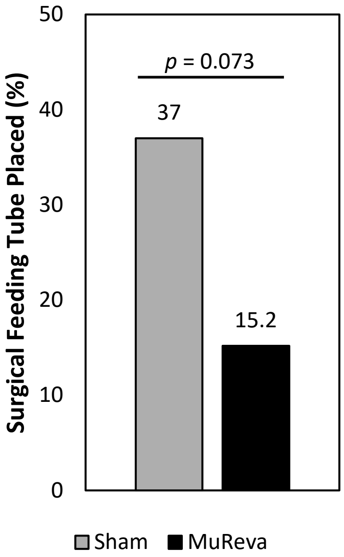


* Excludes participants with a surgical feeding tube at baseline.
